# Nowcasting CoVID-19 Deaths in England by Age and Region

**DOI:** 10.1101/2020.09.15.20194209

**Authors:** Shaun Seaman, Pantelis Samartsidis, Meaghan Kall, Daniela De Angelis

**Affiliations:** MRC Biostatistics Unit, University of Cambridge Institute of Public Health, Cambridge Biomedical Campus, CB2 0SR, UK; National Infection Service, Public Health England, 61 Colindale Avenue, NW9 5HT, UK

## Abstract

Understanding the trajectory of the daily numbers of deaths in people with CoVID-19 is essential to decisions on the response to the CoVID-19 pandemic. Estimating this trajectory from data on numbers of deaths is complicated by the delay between deaths occurring and their being reported to the authorities. In England, Public Health England receives death reports from a number of sources and the reporting delay is typically several days, but can be several weeks. Delayed reporting results in considerable uncertainty about the number of deaths that occurred on the most recent days. In this article, we estimate the number of deaths per day in each of five age strata within seven English regions. We use a Bayesian hierarchical model that involves a submodel for the number of deaths per day and a submodel for the reporting delay distribution. This model accounts for reporting-day effects and longer-term changes over time in the delay distribution. We show how the model can be fitted in a computationally efficient way when the delay distribution is same in multiple strata, e.g. over a wide range of ages.

## 1 Introduction

CoVID-19 appeared in the UK in early 2020, with its first case, imported from China, reported on the 30th January, and the first CoVID-linked death announced on the 6th March. Over the last few months the pandemic has spread in the community, hospitals and care homes. As at 5th September 2020, around 300,000 cases have been lab-confirmed and of these over 40,000 have died. The lock-down introduced on the 23rd of March has drastically reduced transmission, although the process of easing it is proving very challenging. Underlying the governmental response is surveillance information on a number of epidemiological indicators. In particular, trends in the number of deaths have been crucial to monitoring the burden of CoVID-19 as well as to informing models reconstructing and predicting the pandemic. However, information on these trends is affected by delayed reporting. While the number of deaths in people with lab-confirmed CoVID-19 infection peaked on 9th April, the number of reports of such deaths peaked 13 days later, on 22nd April. This is because deaths are rarely reported to the authorities on the day they occur, and some deaths take weeks to be reported. This results in right-truncated data: deaths occurring on day *t* are only observed if their reporting delay *d* is such that day *t* + *d* is earlier than the day up to which data are available [3]. This right-truncation makes it difficult to establish the true pattern of the numbers of deaths occurring over time and, in particular, to identify quickly any increase in the most recent days. The process of estimating the number of deaths that occurred on each day from the numbers of deaths that have so far been reported is referred to as ‘nowcasting’. In this article, we nowcast the number of deaths in people with CoVID-19 both for the whole of England and separately for seven English regions, and we break down these numbers by five age strata: 0–44 years, 45–54 years, 55–64 years, 65–74 years and ≥ 75 years. To produce these nowcasts, we jointly model the distributions of reporting delays, the observed numbers of reported deaths and the actual numbers of deaths in each of the age strata within a region.

We use data on deaths in people with a laboratory-confirmed CoVID-19 diagnosis reported to Public Health England (PHE). These reports come from three sources: National Health Service England (NHSE), the Demographics Batch Service (DBS) and Health Protection Teams (HPT). NHSE reports include deaths occurring in hospitals notified by NHS trusts through the CoVID-19 Patient Notification System and reported daily to PHE. DBS deaths result from the daily tracing of CoVID-19 positive tests reported to PHE through the Second Generation Surveillance System in the NHS Spine to identify individuals who might have died in the previous 24 hours in any setting. HPT data include deaths notified by PHE HPT during outbreak management, primarily in non-hospital settings (e.g. care homes). These three data sources are not independent; duplications are identified daily and only the earliest report date is considered.

We consider data from 22nd March up to 29th June. We chose 22nd March because the first reports from DBS became available on 21st March and some of the deaths reported on 21st would have been reported earlier had DBS been used earlier. At 29th June there were a total of 37529 confirmed reports of deaths that occurred between 22nd March and 29th June. Figure 1a shows the number of deaths that occurred on each day and had been reported by 29th June. The effect of reporting delay is particularly evident in the last three days: while the numbers of deaths per day in the week up to 25th June were between 50 and 100, the numbers for 27th, 28th and 29th June were 12, 9 and 0. Figure 1b shows the number of deaths that were reported on each day. It can be seen that the number of reports tends to be lower on Sundays and Mondays, probably due to some staff not working during the weekend. This is particularly so in more recent weeks. In addition, the number of reports over a single Tuesday-to-Saturday period varies considerably from day to day. It is particularly low, for example, on 26th May.

**Figure 1:**
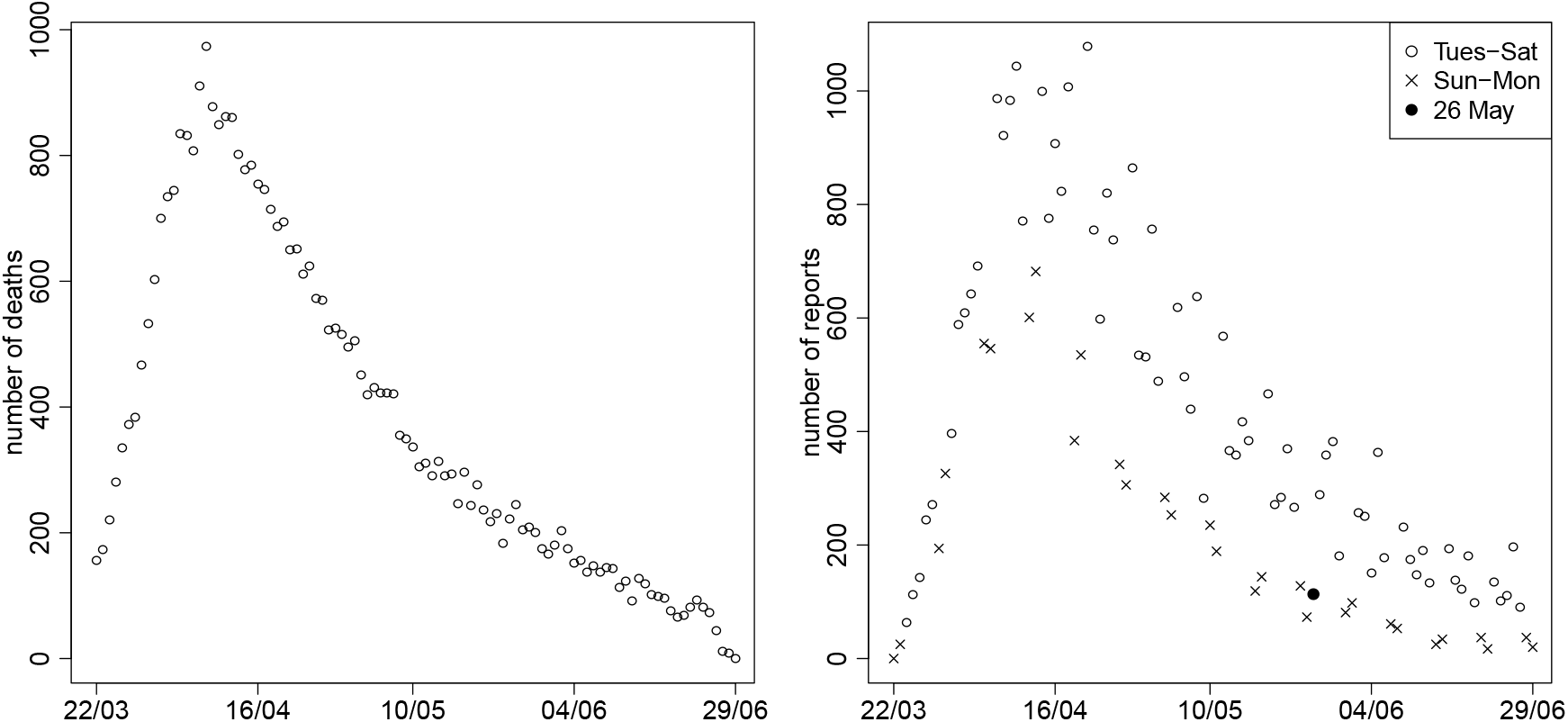
Numbers of reported deaths by a) date of death and b) date of report

Figure 2 shows a simple estimate (see [3] for method) of the distribution of reporting delay by age strata, calculated using data from the whole of England and assuming that the delay distribution is the same for all times of death. This distribution is conditional on the delay being at most 42 days. Forty-two days was chosen because only 61 of the 37529 reported deaths had delays longer than six weeks. Compared to the three strata of individuals aged 45–74, the delays tend to be slightly longer on average in the ≥ 75 years stratum and even more so in the 0–44 years stratum.

**Figure 2:**
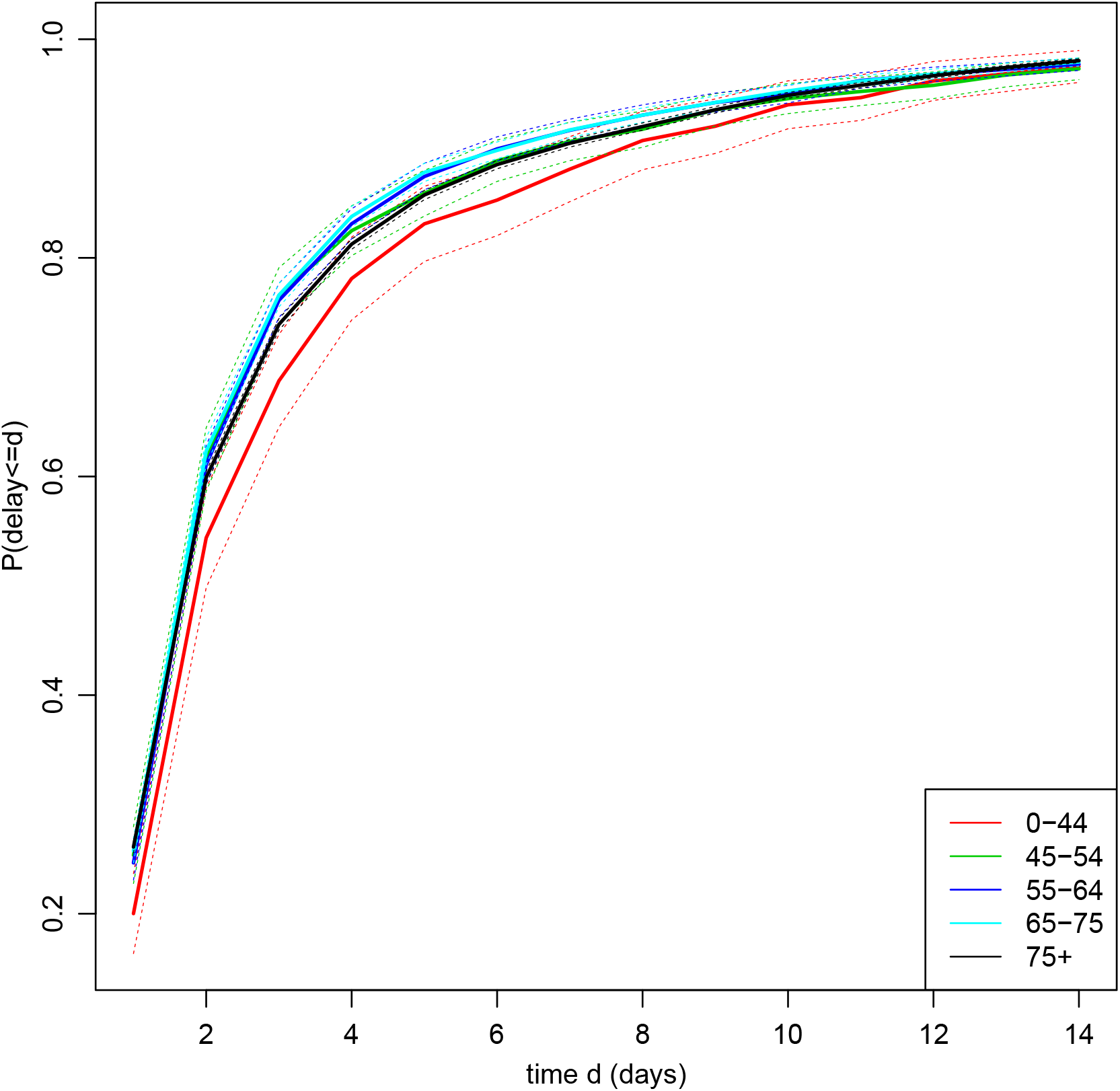
Estimated reporting delay distribution by age stratum. Dotted lines represent 95% CIs. For ease of viewing, only the first 14 days are shown. The probability that the delay is ≤ 14 days is 0.97 for all five strata.

A simple way to adjust the observed figures for reporting delay is to use the estimates in Figure 2 (or corresponding estimates based on data from a single region) to weight up the numbers of deaths reported so far [3]. For example, of the 12 deaths that occurred on 27th June and had been reported by 29th June, nine were in the ≥ 75 strata. The estimated proportion of deaths reported within two days in this strata is 0.60. Therefore, the actual number of deaths in this strata can be estimated as 9/0.60 = 15. However, this estimate does not account for any dependence of the delay distribution on the day of death. Delays may length or shorten over calendar time and may depend (as Figure 1b suggests) on the weekday on which the death occurred. Also, while the numbers of deaths occurring on each day are likely to change smoothly over calendar time, this estimate makes no use of information about the numbers of deaths that occurred on the preceding and succeeding days, with the result that it can be very different from the estimates for those days. In addition, simple confidence interval calculations [3] underestimate uncertainty, unless the delay distribution really is the same for all times of death.

The problem of adjusting numbers of reported events to account for reporting delay arises in several contexts, including insurance claims [6] and epidemiology. Much work was done in the 1980’s in the context of HIV/AIDS, e.g. [2, 3]. Adjustment is complicated by weekday reporting effects, changes over calendar time in the reporting delay distribution, and the existence of days for which no deaths have yet been reported. Stoner and Economou (henceforth S&E) (2019) [10] provide a review of some of the more recent literature on delayed reporting. A number of approaches have been used for nowcasting CoVID-19 in England. Bird and Nielsen (2020) [1] use a modification of the chain-ladder approach employed in general insurance. To allow for change over time in the delay distribution, they only use data from the previous seven days, estimating the numbers of deaths that will be reported within seven days of death. S&E (2020) [9] use a hierarchical Bayesian

The general approach of S&E has several advantages. It allows flexible modelling of weekly and longer-term changes in the reporting delay distribution. It accounts for overdispersion in the delay distribution, which ensures that variation in this distribution which cannot be explained by such changes does not lead to the estimates of the numbers of deaths having excessive variance [10]. By modelling the expected number of deaths per day in each stratum as a smooth function of time, it allows the estimate of the number of deaths that occurred on one day to borrow information about the numbers of deaths that occurred on the preceding and succeeding days. Also, information about the delay distribution and pattern of the epidemic in one strata can be borrowed to inform those quantities in the other strata. Uncertainty can be quantified by posterior credible intervals.

In this article, we build on S&E’s hierarchical model to nowcast in each region, with stratification by age group. Our model improves on theirs in a number of ways. First, we model weekday reporting effects in a more realistic fashion. As mentioned above, fewer deaths are reported on Sundays and Mondays, and the strength of this weekend effect changes over time. Second, we account for correlation between the numbers of reports made on the same day about individuals who died on different days. If, for example, the data suggests that an unusually low proportion of those deaths that occurred between two and seven days ago have been reported today, it is likely that few of the deaths that occurred yesterday and today will have yet been reported. Third, we describe a computationally efficient way to constrain the delay distribution to be the same in multiple strata. This is particularly useful if the delay distribution can be expected to be the same across a wide range of ages. We also illustrate how to use the model to calculate a posterior probability that the number of deaths has been rising in recent days.

## 2 Methods

Denote the first day for which data on reporting delays will be used as day 0 and the most recent day for which data are available as day *T*.

We shall model deaths that are reported within *D* days. For the CoVID-19 data, we set *D* = 42, because, as noted in Section 1, very few deaths were reported with delays greater than six weeks. Let *D_t_* denote the longest delay that we could have observed in individuals who died on day t, excluding any individuals with delays greater than *D* days. So, *D_t_* = *D* if *t* ≤ *T* − *D* and *D_t_* = *T* − *t* otherwise. Let *Z_std_* denote the number of deaths that occurred on day *t* (*t* = 0,…, *T*) in age stratum *s* (*s* = 1,…, *S*) and are reported with a delay of *d* days (*d* = 0,…, *D*). We observe *Z_std_* when *d* ≤ *D_t_*; otherwise *Z_std_* is unobserved. We used *S* = 5 strata: 0–44 (*s* = 1), 45–54 (*s* = 2), 55–64 (*s* = 3), 65–74 (*s* = 4) and ≥ 75 (*s* = 5) years. Let 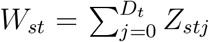 be the observed number of deaths reported in stratum *s* by time *T* among individuals who died on day *t*. Let 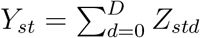 be the number of deaths that occurred on day *t* (*t* = 0,…, *T*) in age stratum s and are reported with a delay of at most *D* days. For *t* = 0,…, *T* − *D, Y_st_* is observed: it equals *W_st_*. For *t* = *T* − *D* + 1,…, *T, Y_st_* is unobserved and we seek to estimate it.

We model the data in a Bayesian framework, by specifying three submodels: one for the marginal distribution of the number of deaths, *Y_st_*, occurring on each day in each stratum; one for the conditional distribution of the observed numbers of reports, *Z_std_*, given the numbers of deaths and the delay distribution; and one for the delay distribution. Figure S1 shows a directed acyclic graph for the model (all figures prefixed by ‘S’ are in the Supplementary Materials). We fit this Bayesian model using a Markov chain Monte Carlo (MCMC) algorithm.

### 2.1 Model for numbers of deaths

The number of deaths, *Y_st_*, that occurred in stratum *s* on day *t* is unobserved for *t* > *T* − *D*. Assume that, for stratum *s* = 1,…, *S* and time *t* = *T* − *D* + 1,…*T*,

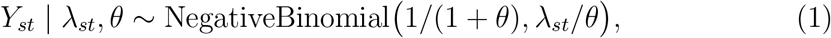

where λ*_s_,_T_*_−_*_D_*_+1_,…, λ*_sT_* and *θ* are unknown parameters. λ*_st_* = *E*(*Y_st_* | λ*_st_*, *θ*) is the expected number of deaths on day *t* in stratum *s*, and Var(*Y_st_* | λ*_st_*, *θ*_s_) = λ*_st_*(1 + *θ*). So, *θ* (*θ* ≥ *θ*) is an overdispersion parameter. In particular, if *θ* = 0, then *Y_st_* | λ*_st_*,*θ* ~ Poisson(λ*_st_*). We shall assume that λ*_st_* changes smoothly over time *t*.

The model described in the last paragraph does not use data on the observed numbers of deaths *Y_st_* that occurred in the period before day *T* − *D* + 1. Such data may be useful, as they indicate whether the number of deaths per day was rising or falling over that period. It could reasonably be expected that the trajectory of the number of deaths per day in the period shortly after day *T* − *D* + 1 would be a continuation of the trajectory immediately before it. To exploit this information, we shall assume that expression (1) also holds for *t* = *T*_0_,…, *T* − D, where *T*_0_ is some constant between 0 and *T* − *D*. We chose T_0_ = *T* − *D* − 13, so that we use the most recent two weeks for which *Y_st_* is observed.

We follow S&E in using restrictive penalised cubic splines to model the assumption that λ*_st_* changes smoothly over time t. Furthermore, we assume that 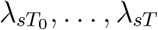 may follow similar trajectories in different strata *s*, while allowing these trajectories to differ. More specifically, log λ*_st_* (*t* = *T*_0_,…, *T*) is taken to be the sum of an unknown stratum-specific intercept term *ι_s_* and two restricted cubic splines, one of which is common to all strata and one which is specific to stratum *s*. That is, 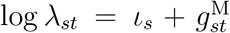, where 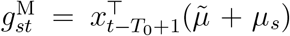 (superscript ‘M’ stands for ‘mortality’). Here, 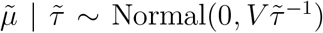 and 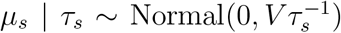, 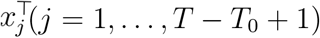 is the *j*th row of the design matrix *X* for the splines, and *V* is the corresponding variance matrix [11, 12]. The unknown parameters 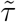 and *τ_s_* determine the smoothness of the splines. If *τ_s_* = ∞ for all *s*, then *μ_s_* = 0, and so λ*_st_* = λ*_t_*, i.e. the patterns of mortality in the five strata are the same. The R function *jagam* [11] can be used to calculate *X* and *V*.

### 2.2 Model for observed number of reports

For stratum s and time t, we model *Z_st_*_0_,…, *Z_stD_* as

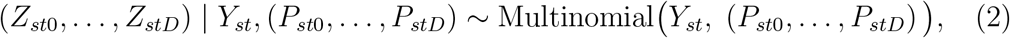

where *P_std_* is the unknown probability that an individual in stratum *s* who died on day *t* is reported with a delay of *d* days.

We shall allow *P_std_* to depend on *t* through both a smooth function of calendar-time and weekday reporting effects: average reporting delays may gradually shorten or lengthen over calendar time and Figure 1 shows deaths are less likely to be reported on Sundays and Mondays. However, even after accounting for these systematic effects, the observed numbers of daily death reports *Z_std_* may be overdispersed compared to a multinomial distribution. As S&E (2019) showed, failure to allow for such overdispersion can lead to overdispersion in the numbers of deaths *Y_st_*, and thus to unnecessarily imprecise nowcasts. So, to allow for overdispersion in *Z_std_*, we assume that, for each *s* and *t*, (*P_st_*_0_,…, *P_stD_*) is drawn from a generalised Dirichlet (GD) distribution [4]. That is, the density of (*P_st_*_0_,…, *P_stD_*) is

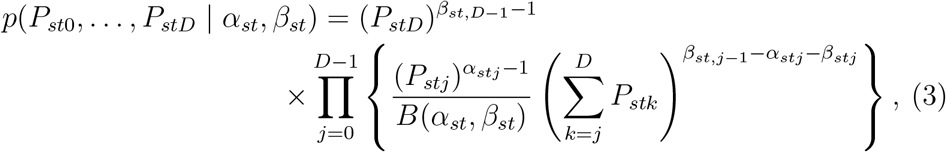

where *B*(.,.) denotes the beta function (which is a ratio of gamma functions) and *α_st_* = (*α_st_*_0_,…, *α_st,D_*_−1_) and *β_st_* = (*β_st_*_0_,…, *β_st,D_*_−1_)) are unknown parameters that will be modelled as functions of *s*, *t* and *d* (note that *β_st,j_*_−1_ = 0 when *j* = 0). The GD distribution is a flexible generalisation of the Dirichlet distribution; it allows, for example, positive correlation between *P_std_* and *P_std_*_′_ for *d ≠* d′, unlike the Dirichlet distribution [4, 10].

The marginal distribution of (*Z_st_*_0_,…, *Z_stD_*) (given *Y_st_*, *α_st_* and *β_st_*) obtained by integrating out (*P_st_*_0_,…, *P_stD_*) is the generalised-Dirichlet-multinomial (GDM) distribution, which has a closed-form expression [4, 10]. This GDM distribution can be factorised as the product over *d* = 0,…,*D* − 1 of the conditional distribution of *Z_std_* given *Z_st_*_0_,…, *Z_st,d_*_−1_, *Y_st_*, *α_st_* and *β_st_*, which is

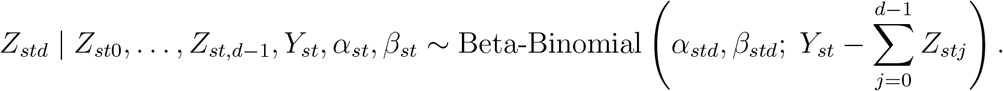

This distribution has probability mass function

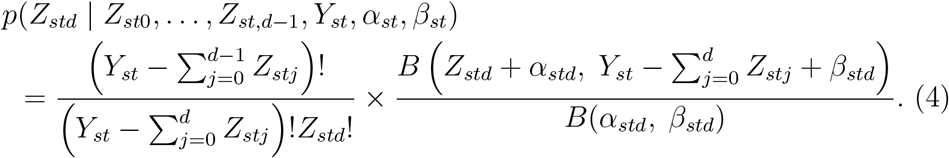

Since *Z_std_* is unobserved for *d* > *D_t_*, we only need include the likelihood contribution of (*Z_st_*_0_,…, *Z_stDt_*), i.e. the product of expression (4) over *d* = 0,…, min(*D_t_*, *D*−1). By integrating out the unknown random variables 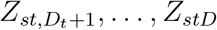 in this way, the Markov chain Monte Carlo (MCMC) algorithm used to fit the model is simplified compared to the algorithm used by S&E, the computational load is reduced, and the mixing of the Markov chain should be improved. Note that the *Z_std_*! term in expression (4) can be ignored when calculating full-conditional distributions, because it does not depend on any unobserved variables.

### 2.3 Model for the delay distribution

We reparameterise the GD distribution in terms of parameters *υ_st_* = (*υ_st_*_0_,…, *υ_st,D_*_−1_) and *ϕ_st_* — (*ϕ_st_*_0_,… *ϕ_st,D_*_−1_), where υ*_std_* = *α_std_/*(*α_std_* + *β_std_*) and *ϕ_t_* = *α_std_* + *β_std_*. Now, 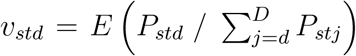 is the expected hazard of a reporting delay of *d* days for an individual in stratum *s* who died on day *t* [4]. The variance of this hazard is

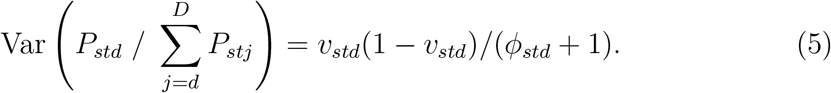

Note, in particular, that as *ϕ_std_* → ∞, this variance tends to zero, which makes the GDM distribution of (*Z_st_*_0_,…, *Z_stD_*) | *Y_st_* reduce to the multinomial distribution of expression (2) with 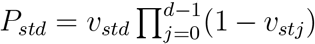. Correspondingly, the beta-binomial distribution of equation (4) reduces to a binomial distribution. Thus, the *ϕ_std_* parameters describe the overdispersion of the numbers of reports relative to a multinomial distribution.

It is easy to show that the expected hazard *υ_std_* can also be written as

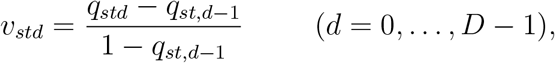

where 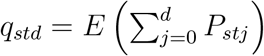 is the expectation of the probability that the delay is at most *d* days for an individual in stratum *s* who died on day *t*.

#### 2.3.1 Weekday effects

We model weekday reporting effects by assuming for *d* = 0, ..., *D* − 1 that

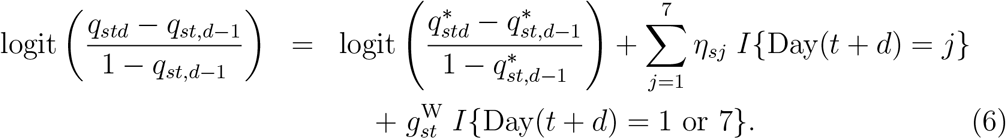

Here, *I*(.) is the indicator function, and Day(*t* + *d*) = 1 if day *t* + *d* is Monday, Day(*t* + *d*) = 2 if day *t* + *d* is Tuesday, and so on. The quantities 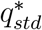 and *η_s_*_1_, *η_s_*_2_, *η_s_*_3_, *η_s_*_4_, *η_s_*_5_, *η_s_*_7_ are unknown parameters; we constrain *η_s_*_6_ = 0 for identifiability (Saturday is our baseline). The function 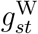 is a smooth function of *t*, which describes how the weekend effect (i.e. the effect of its being Sunday or Monday) changes over calendar time. As we did earlier with 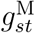, we assume 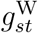 is the sum of two restricted cubic splines, one common to all strata and one specific to stratum *s*. Equation (6) means that, given the delay is at least *d* days, the odds of a death on day *t* being reported with a delay of *d* is 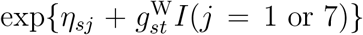 times greater when day *t* + *d* is weekday *j* than it would be if day *t* + *d* were Saturday.

#### 2.3.2 Calendar-time effects

To model calendar-time effects, we assume for *d* = 0, ..., *D* − 1 that

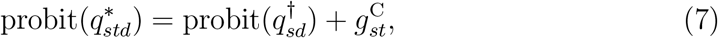

where 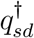 is an unknown parameter and 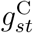 is a smooth function of *t*. As with 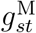 and 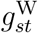, we assume 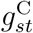 is the sum of two restricted cubic splines, one common to all strata and one specific to stratum *s*.

To ensure that 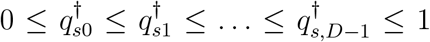, and hence 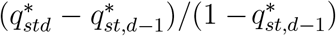 in equation (6) lies between 0 and 1, we reparameterise 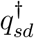 as 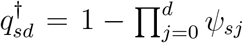, where *ψ_s_*_0_,…, *ψ*_s,D-1_ take any values between 0 and 1. Note that if 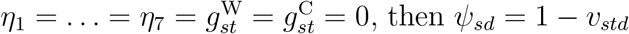, then *ψ_sd_* =1 − *υ_std_* and 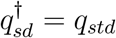.

In Section 4, we contrast our approach to modelling weekday and calendar-time reporting effects with the approach used by S&E.

#### 2.3.3 Reporting-day random effects

The weekday effects *η_sj_* and 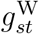 are reporting-day effects: they reflect the phenomenon that deaths may be more likely to be reported on certain days of the week. In addition to these systematic reporting-day effects, there may be non-systematic reporting-day effects. That is, there may be individual days on which the numbers of reports is unpredictably higher or lower than would be expected given the day of the week and the number of deaths that have occurred in recent days. Figure 1b suggests this is true, for example, of 26th May. These reporting-day effects would alter the expected hazard *υ_std_* on day *t* + *d* for delays of all lengths *d* = 0,…, *D* − 1, and hence induce correlations between *P_std_*, *P_s,t_*_+1,_*_d_*_−1_, *P_s,t_*_+2,_*_d_*_−2_, *P_s,t_*_+3,_*_d_*_−3_,…. Such correlations are not reflected by the GD distribution of equation (3), which instead induces correlations between *P_st_*_0_, *P_st_*_1_, *P_st_*_2_, *P_st_*_3_,…. To allow for these non-systematic reporting-day effects, we add a random effect *δ_t_*_+_*_d_* to equation (6), i.e. we change (6) to

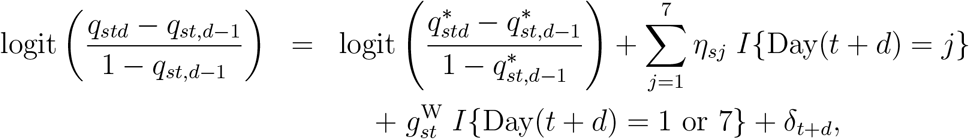

where *δ*_0_,…,*δ_T_* are i.i.d. Normal(0, *ω*^2^) and *ω* is an unknown parameter.

#### 2.3.4 Strata with a common delay distribution

The model described so far assumes the delay distribution for individuals who die on day *t* is different in each stratum *s*. For the CoVID-19 data, it is reasonable to assume the delay distributions are equal in strata *s* = 2, 3 and 4, i.e. individuals aged 45−74. We considered two alternative adaptations of the model to impose the constraint that the delay distribution is the same in some group 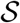 of strata (where 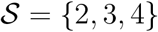 for the CoVID-19 data).

The first adapted model sets the parameters and splines *η_sj_*, *ψ_sd_*, 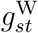 and 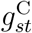 that determine the delay distribution in stratum s to be equal for all 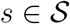. We denote the common values by a dot, so *η_sj_* = *η_•j_*, *ψ_sd_* = *ψ*_•_*_d_*, 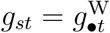 and 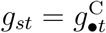. Now *υ_std_*, the expected hazard of reporting on day *t* + *d* for an individual who died on day *t*, is the same for strata 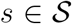. If, in addition, *ϕ_std_* = *ϕ*_•_*_td_* for all 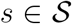, then 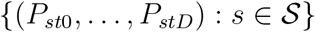 have the same GD distribution.

The second adapted model assumes *P_std_* = *P*_•_*_td_* for all 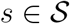, i.e. delay distributions 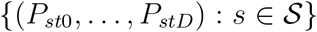 are exactly equal. Let 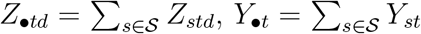 and 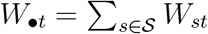. We show in the Appendix that when *P_std_* = *P*_•_*_td_*,

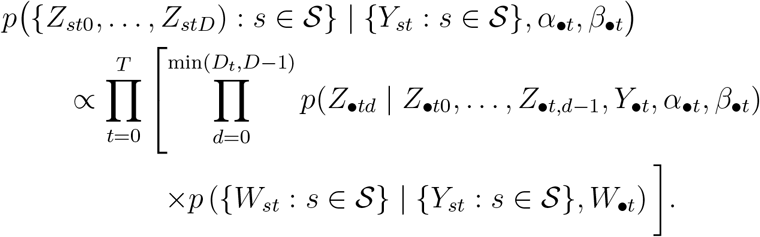

Assuming (*P*_•_*_t_*_0_,…, *P*_•_*_tD_*) is drawn from a GD distribution, we have (as in expression (4)),

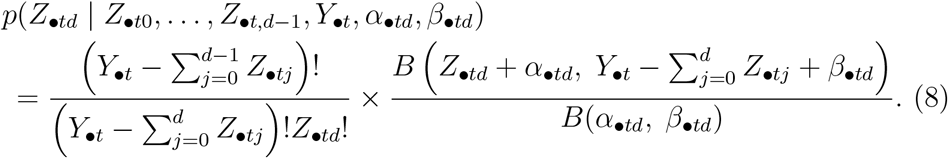

Furthermore, conditional on 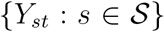 and 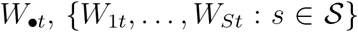 has a multivariate hypergeometric distribution:

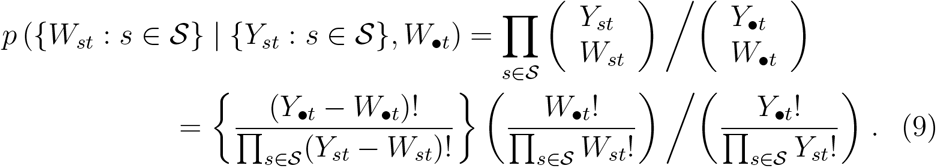

So, in this second adapted model, for each of *t* = 0, …,*T* and each of *d* = 0,…, min(*D_t_*, *D* 2212 1), the 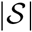 likelihood contributions of expression (4) are replaced by the single likelihood contribution of expression (8), and for each of *t* = 0,…, *T*, there is the additional single likelihood contribution of expression (9).

Compared to the first adapted model, this second model has the advantage of involving less computation. This is because each time *η*_•_*_j_*, *ψ*_•_*_d_*, *δ_j_* or a parameter of the splines underlying 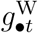 and 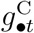 is updated in the MCMC algorithm the single expression (8) is evaluated, rather than the 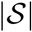 expressions (4). The only time expression (9) is evaluated is when *Y_st_* is updated. Note that the 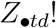 and 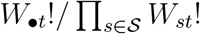 terms in expressions (8) and (9), respectively, can be ignored when calculating full-conditional distributions, because they are functions only of observed variables.

### 2.4 Priors and model fitting

The intercept term *ι_s_* in the model for log λ_st_ was assigned a non-informative normal prior with mean zero and variance 100. For the priors on the precision parameters 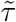 and *τ_s_* of the splines for 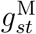 we used the same inverse-gamma with shape and rate 0.5 that S&E (2020) used. We also used this prior for the corresponding precision parameters of the splines for 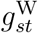 and 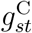. The prior on the dispersion parameter *θ* of the negative binomial distribution for *Y_st_* was an exponential distribution with rate 2. This assigns considerable mass to values close to zero (no overdispersion), while still allowing considerable overdispersion.

For the delay distribution, we used a non-informative uniform prior on *ψ_sd_* and, following S&E (2019), used an exponential distribution with rate 0.01 for the prior on *ϕ_d_*. Weekday effects *η_sj_* were given non-informative normal priors with mean zero and variance 100. The prior on the precision, *ω*^−2^, of the reporting-day random effects was a gamma distribution with shape 1 and rate 0.005. This implies a prior mean of 0.125 on the standard deviation *ω*, with 90% prior credible interval (0.04, 0.31). The value *ω* = 0.125 corresponds to 95% of the odds ratios exp(*δ_t_*_+_*_d_*) lying between 0.78 and 1.28; *ω* = 0.31 corresponds to them lying between 0.54 and 1.84.

The model was fitted using a MCMC algorithm, implemented in R using the NIMBLE package [5]. R code provided by S&E (2020) was used as a template for our code. Our R code is provided in the supplementary materials. The unknown random variables that appear (and are updated) in our MCMC algorithm are: the intercepts *ι_s_*; the weekday effects *η_s_*_1_,…, *η_s_*_7_; the parameters *ψ_s_*_0_,…, *ψ_s,D_*_−1_ of the delay distribution; the numbers of deaths *Y_s,T_*_−_*_D_*_+1_,…, *Y_sT_*; the reporting-day random effects *δ*_0_,…, *δ_T_* and their standard deviation *ω*; the parameters 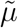, *μ_s_*, 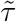 and *τ_s_* of the splines underlying 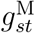; and the corresponding parameters of the splines underlying 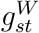 and 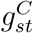. See Figure S1 for a directed acyclic graph of the model. We generated 300,000 iterations, of which the first 50,000 were discarded as burn-in.

## 3 Results

In this section, we describe results from analysing data from the whole of England. Nowcasts produced by analysing the data from each of the seven NHS regions separately are provided in the Supplementary Materials. We take the delay distribution to be the same in the three strata aged 45-74, using the second adapted model described in Section 2.3.4.

For these data, we made two modifications to the model described in Section 2. First, as Figure 2 shows, only 3% of delays are longer than 14 days. We grouped these into weeks. That is, delays of 15–21 days were grouped together, as were 22–28, 29–35 and 36–42 days. We also grouped delays of zero days and one day, because only 445 (4.6%) of the 9719 delays of zero or one day were equal to zero. Second, when fitting the model by MCMC, we struggled to achieve good mixing of 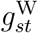 and 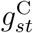, the weekend and calendar-time effects. This task may have been made more difficult by the small number of age groups (3) and large imbalance in the numbers of deaths in these groups (461, 100042 and 27020 in the 0–44, 45–74 and ≥ 75 groups, respectively; we excluded six deaths in people with unknown age). To improve the mixing of 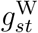 and 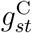, we removed, for both the weekend and the calendar-time effects, the spline specific to the ≥ 75 group, leaving one spline for the effect in the ≥ 75 stratum, one for the difference between the effects in the 0–44 and ≥ 75 strata, and one for the difference between the 45–74 and ≥ 75 groups.

### Weekday reporting effects

Figure 3 shows the posterior distribution of the weekday effects *η_sj_* in the ≥ 75 age stratum. As expected, the hazard of reporting is lower on Sundays and Mondays than on other days. It is also somewhat higher on Fridays than on the remaining days. Figure 4 shows the posterior mean and pointwise 95% posterior credible interval (CI) of 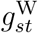, the change in the weekend reporting effect over calendar time. The change is substantial: the posterior mean of 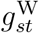 decreases from around 0.7 at the end of March to around −0.9 at the end of June. Since the posterior mean of *η*_57_ is −1.2, this means that the odds ratio of reporting on a Sunday relative to a Saturday is estimated to be around exp(−1.2 + 0.7) = 0.6 in late March but around exp(−1.2 − 0.9) = 0.12 in late June. Figures S2–S5 show the corresponding results for the 0–44 and 45–74 age groups. These are similar to those in the ≥ 75 stratum, except that the credible intervals for the 0–44 stratum are wider, reflecting the smaller numbers of deaths, and hence less information, in this stratum.

**Figure 3:**
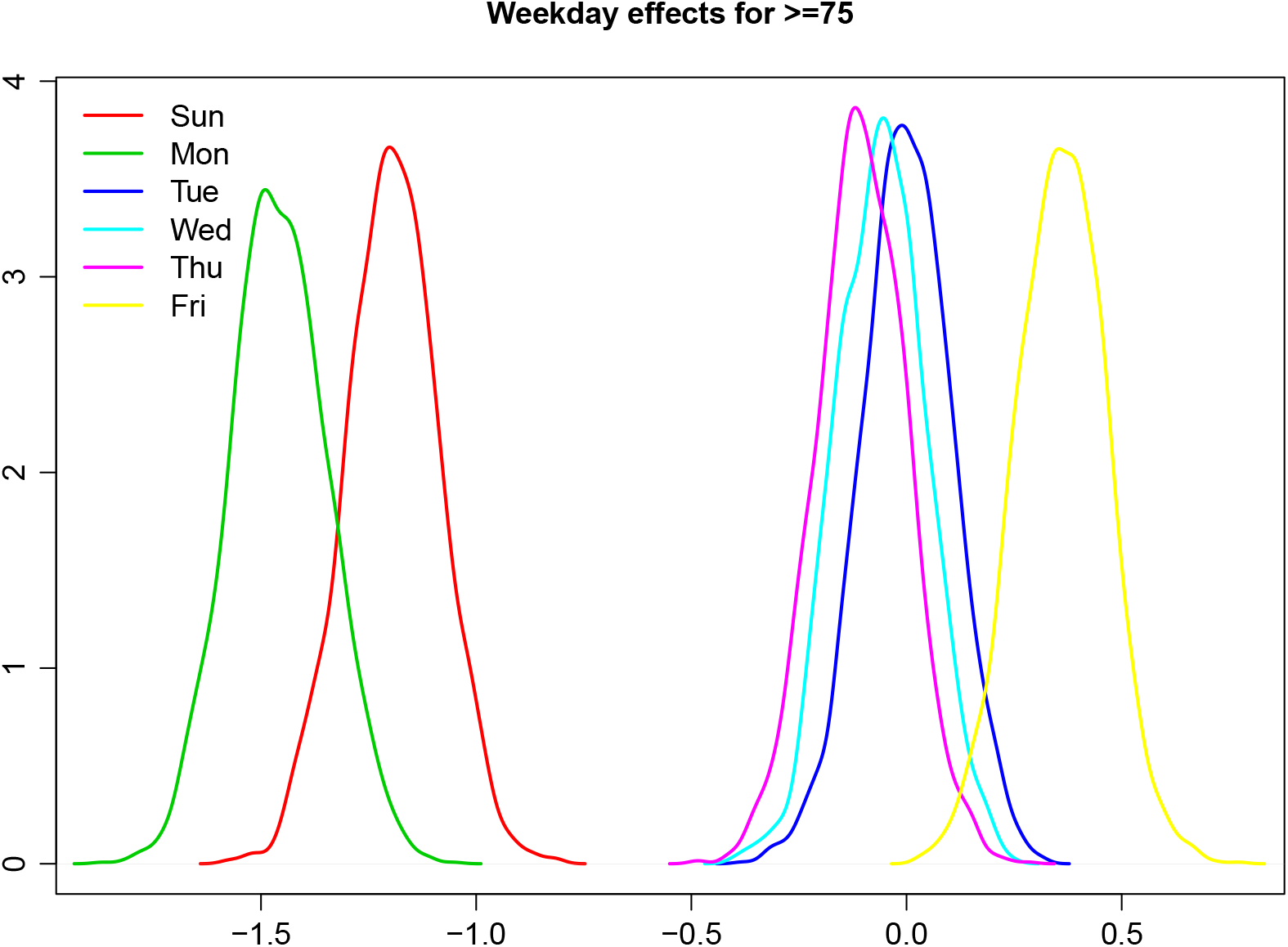
Posterior distributions of day of the week effects (*η_j_*) in the ≥ 75 age stratum

**Figure 4:**
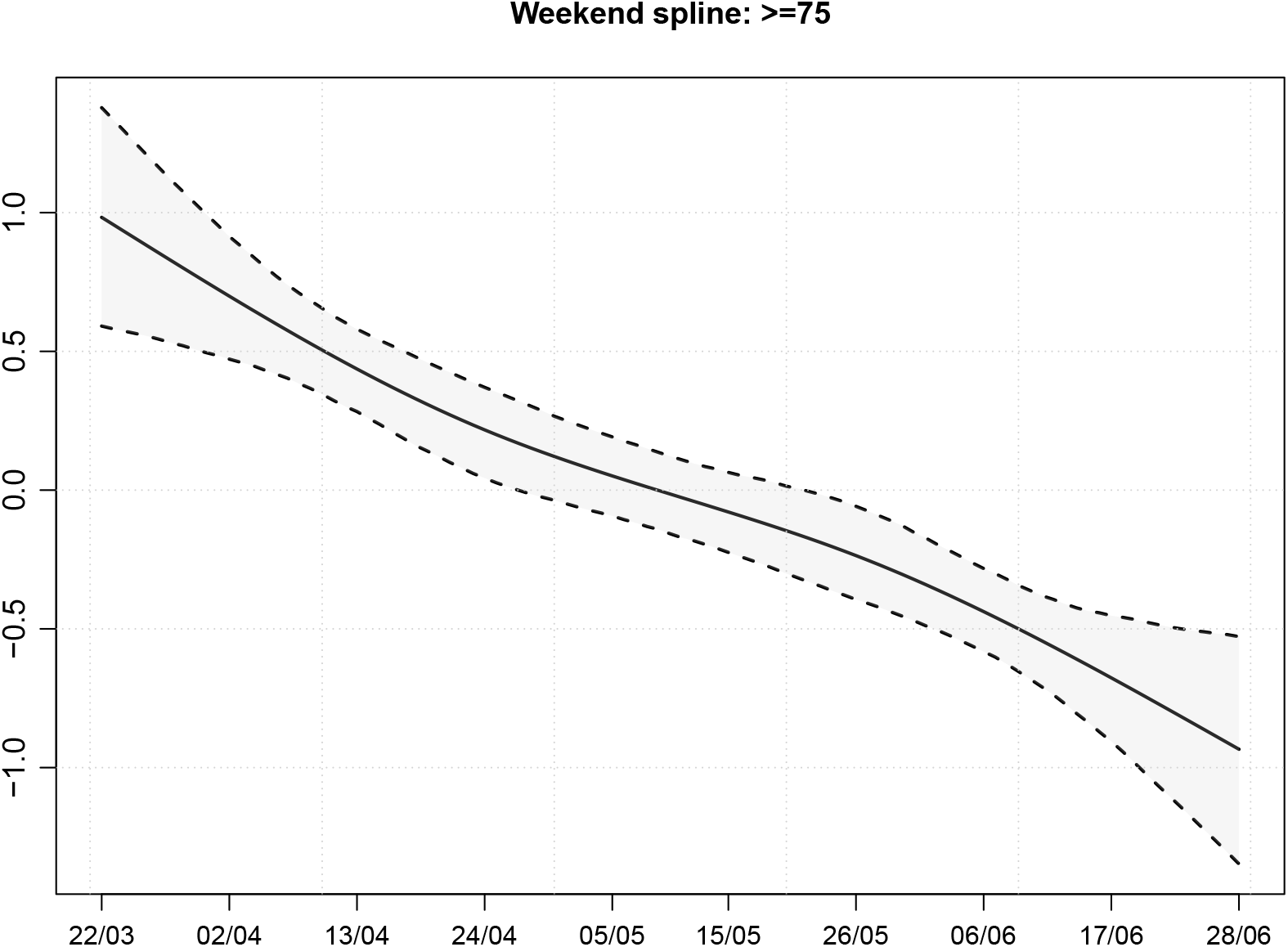
Posterior mean and 95% CI of the change in the weekend effect 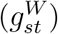 in the ≥ 75 age stratum

### Calendar-time reporting effects

Figure 5 shows the posterior mean and pointwise 95% posterior CI of the calendar-time effect on the delay distribution in the ≥ 75 age stratum. The reporting delays become shorter between 22nd March and the middle of April (the peak of the epidemic), and then lengthen again. In the most recent period, they appear to be shortening again. The pattern is similar in the 45–74 group (see Figure S7). In the 0–44 stratum, the delays also shorten between 22nd March and mid-April, but do not lengthen again after mid-April (Figure S6).

**Figure 5:**
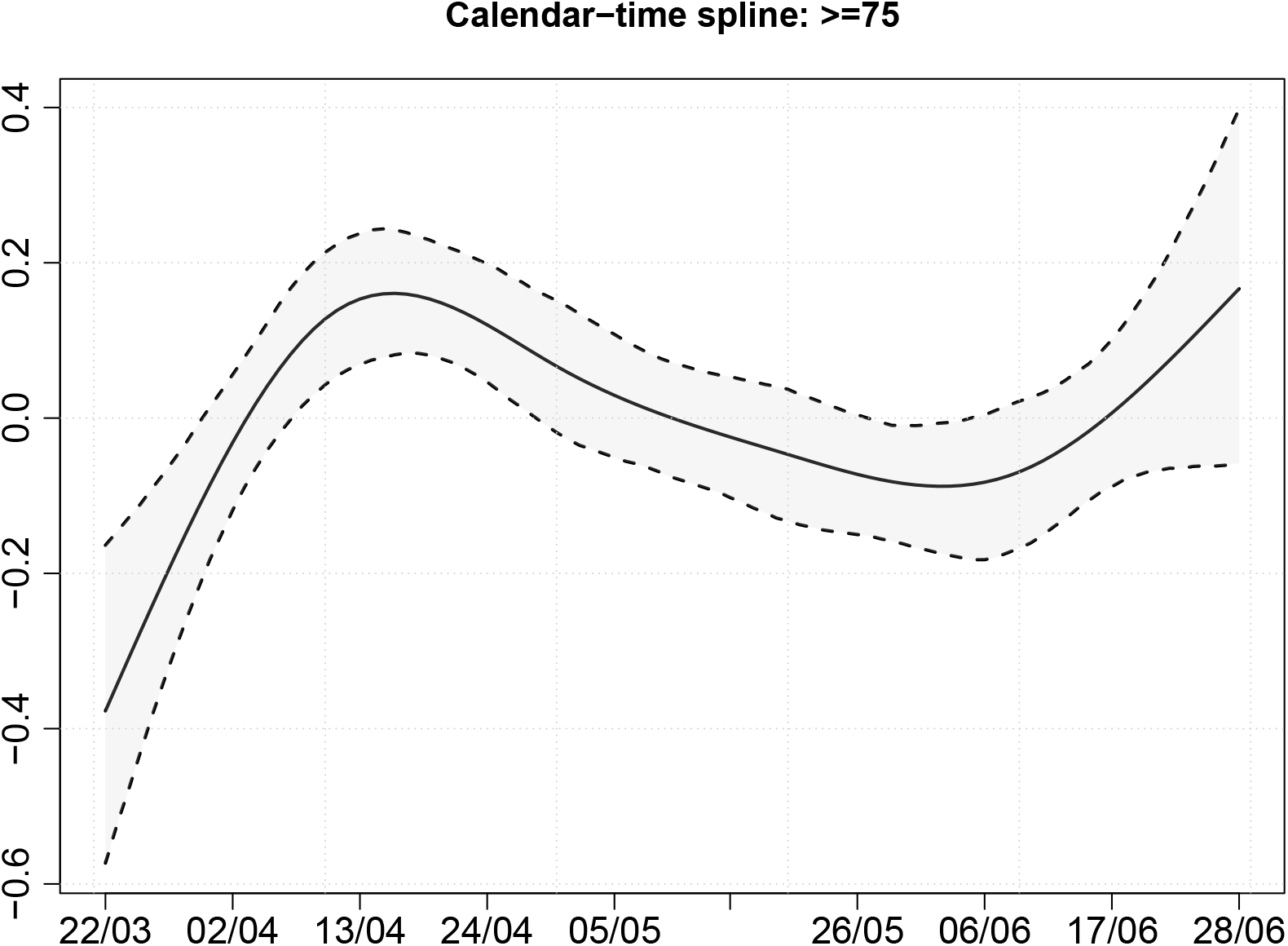
Posterior mean and 95% CI of the calendar-time effect 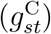 in the ≥ 75 age stratum

### Reporting day random effects

Figure 6 shows the posterior distributions of the reporting-day random effects. The largest random effect is for 26th May; this has a posterior mean of approximately −1. That it is large and negative is unsurprising in view of Figure 1b. There we see that the number of reports on Tuesday 26th May is much lower than the numbers on each of the preceding Tuesday to Saturday and following Wednesday to Friday. Although it is similar to the numbers on the preceding two days, these are Sunday and Monday, when the number of reports is expected to be low. The posterior mean of *ω*, the standard deviation of the random effects, is 0.23, with 95% posterior CI (0.19, 0.29).

**Figure 6:**
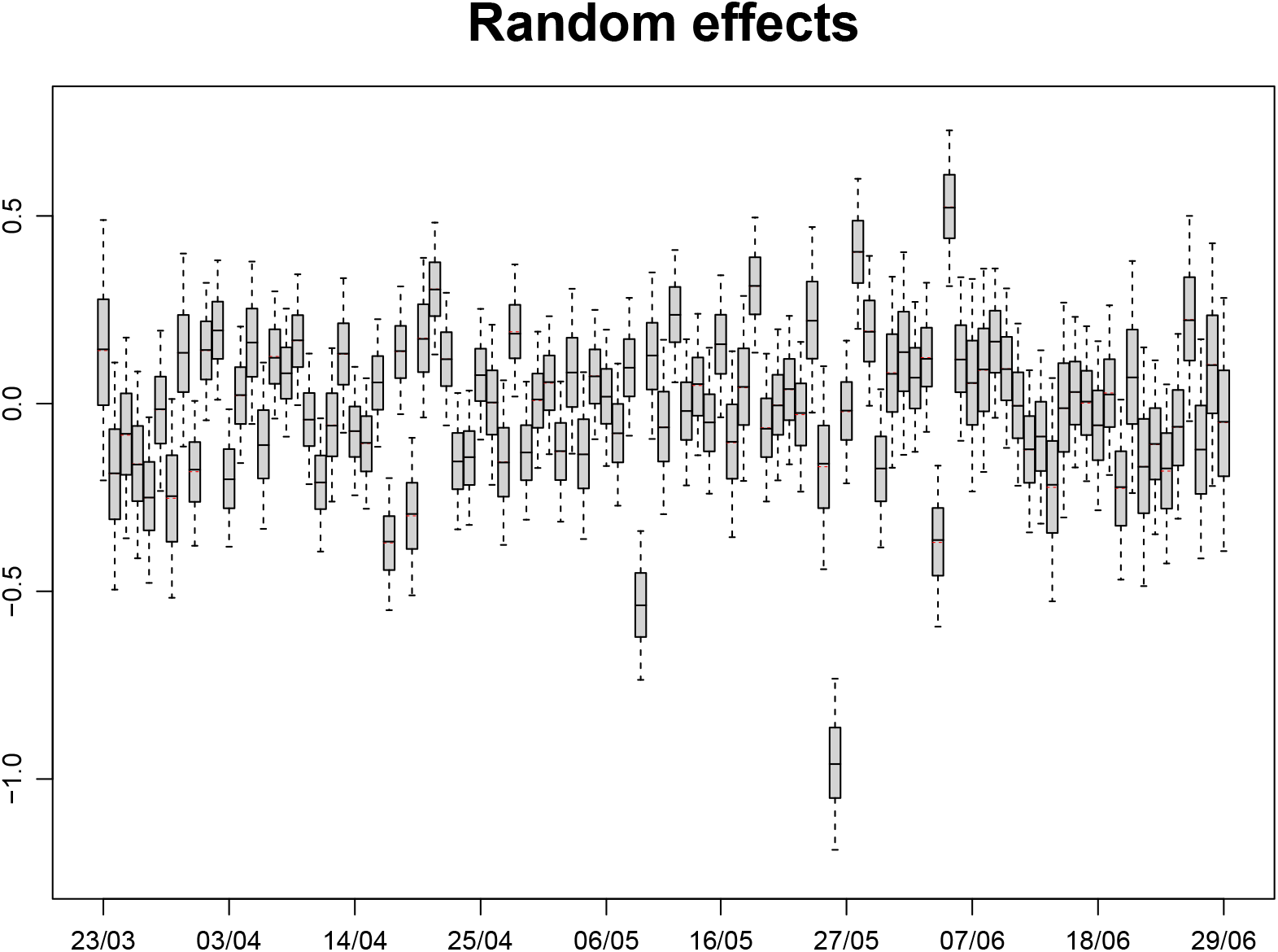
Posterior distributions of reporting-day random effects (*δ_t_*_+_*_d_*). Boxes show posterior median and quartiles, red bars show posterior means, and whiskers show 95% credible intervals.

Dispersion parameters *ϕ_d_* and *θ*

The standard deviation of the reporting hazard 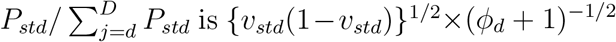 (equation (5)). Figure S8 shows the posterior distribution of (*ϕ_d_* + 1)^−1/2^ for each delay *d*. The posterior means of (*ϕ_d_* + 1)^−1/2^ are between 0.10 and 0.15 for 1 ≤ *d* ≤ 7, and between 0.25 and 0.50 for 7 ≤ *d* ≤ 14. The posterior 95% CIs of (*ϕ_d_* + 1)^−1/2^ for *d* = 1,…, 14 do not include values much lower than 0.1. This suggests that a simpler model that replaced the GDM distribution for (*Z_st_*_0_,…, *Z_stD_*) given *Y_st_* with a multinomial distribution would be inadequate for these data, as such a model would effectively set (*ϕ_d_* + 1)^−1/2^ = 0.

The posterior mean of *θ*, the overdispersion parameter for *Y_st_*, is 0.30, with 95% posterior CI (0.08, 0.55). This suggests mild Poisson overdispersion: the variance of *Y_st_* is around 1.3 times as large as its expectation.

### Nowcasts

Figure 7 shows the nowcasts for the five age strata. For ease of viewing, we show only the most recent 21 days (see Figures S9–13 for the whole period from March 31st). In the ≥ 75 stratum, nine and eight deaths occurred on 27th and 28th June, respectively, and were reported by 29th June. The corresponding estimated numbers of deaths on those two days are 63 and 66, respectively. Hence, it is estimated that only 9/63 = 14% of the deaths that occurred on Saturday 27th June had been reported within two days, and only 12% of deaths that occurred on Sunday 28th June had been reported within one day. These numbers may seem low, considering that Figure 2 shows that, on average, around 26% of deaths in this stratum are reported within one day and around 60% are reported within two days. There are likely to be two reasons for this. First, reporting is much lower on Sundays and Mondays than during the rest of the week, especially in the most recent period. Second, the estimated numbers of deaths for the days before 27th June were all above 60, and it is unlikely that the number of deaths would suddenly drop far below 60. The 95% posterior CIs for these two days are particularly wide, extending from around 45 to 90 deaths.

**Figure 7:**
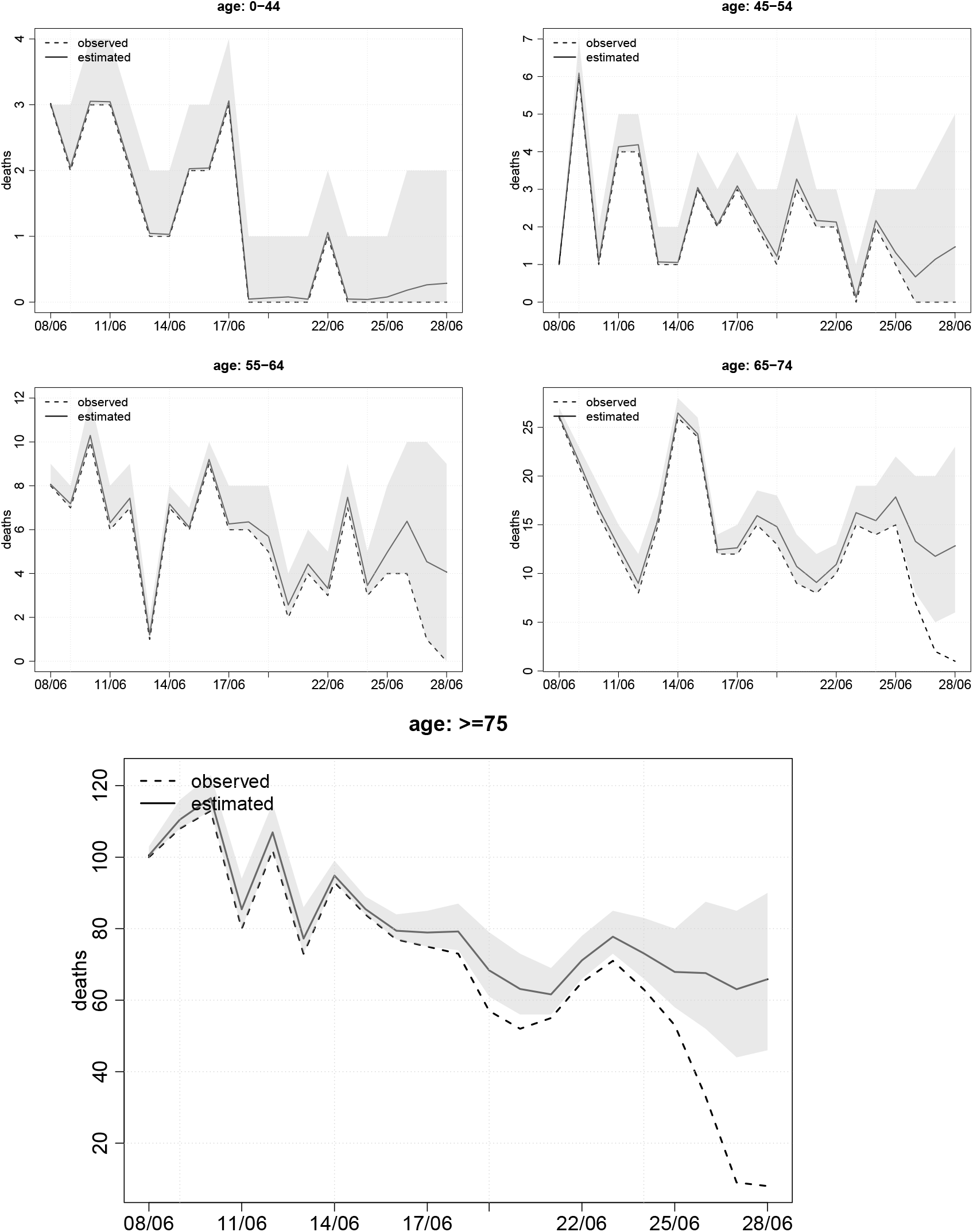
Estimates of the numbers of deaths occurring on each day (solid line), with corresponding numbers of deaths reported by 29th June (broken line). Posterior 95% CIs are shown by shaded region.

### Probability of increase in deaths in recent days

Finally, it is useful to be able to detect early signs of an increasing trend in the number of deaths in recent days. Figure 8 shows, for each of the strata, the posterior probability that the number of deaths in the most recent *x* days was greater than the number in the preceding *x* days (*x* = 1,… 7). There is no convincing evidence of an increase in any of the strata. However, many of the probabilities are in the region of 0.5, meaning that deaths are as likely to be increasing as they are to be decreasing or remaining stable.

**Figure 8:**
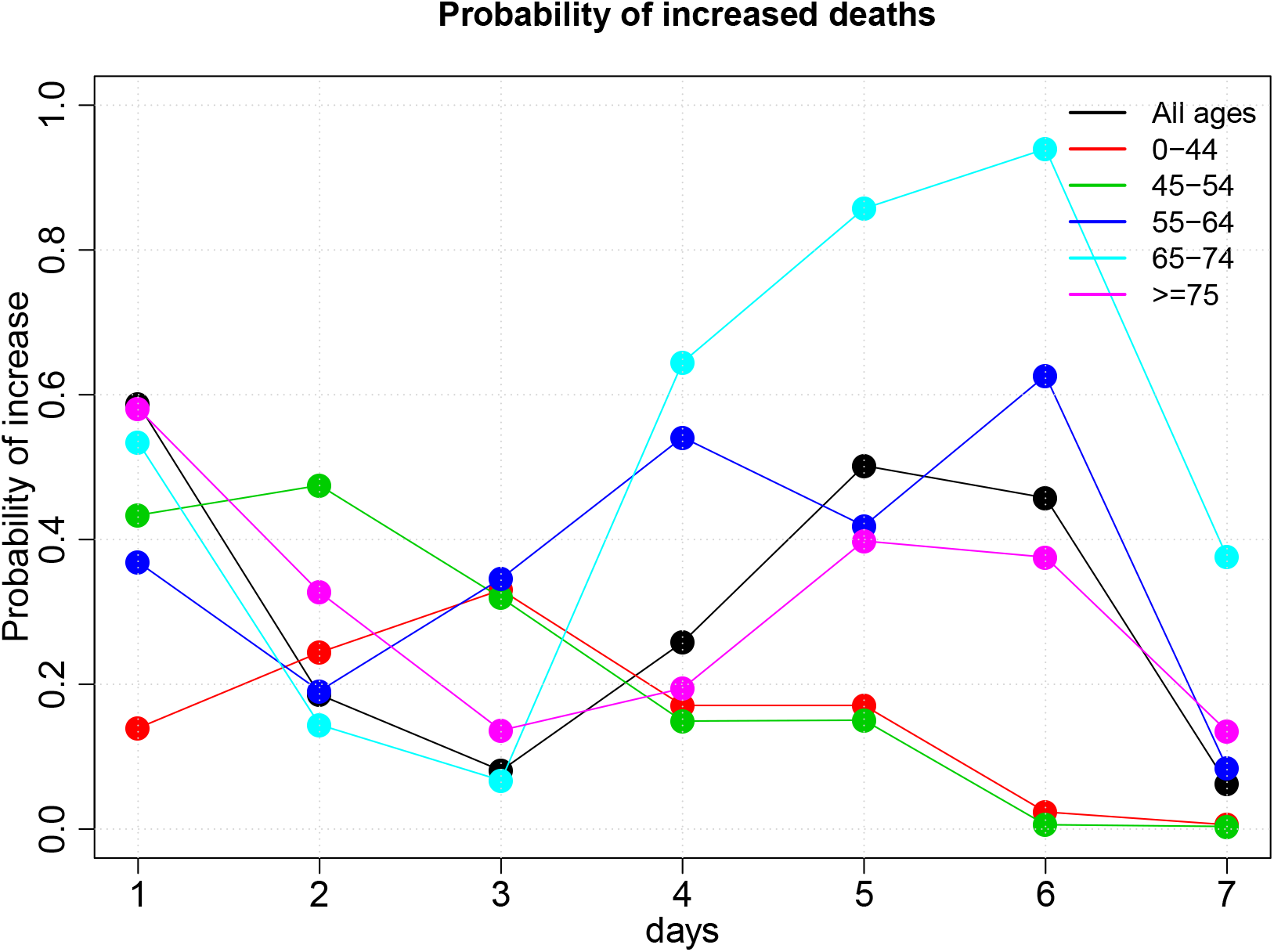
Posterior probabilities that the number of deaths in the most recent *x* days was greater than the number in the preceding *x* days (*x* = 1,… 7)

## 4 Discussion

Timely monitoring of the number of CoVID-19-related deaths relies on methods to adjust the observed data for reporting delay. These methods need to be flexible enough to deal with the peculiar reporting patterns of these data and the way these patterns change over time. Here we built on the approach of S&E and derived age-specific delay-adjusted estimates of the number of deaths occurring in each of the seven English NHS regions.

The modelling of weekday and calendar-time effects described in Sections 2.3.1 and 2.3.2 is different from the way that S&E (2019, 2020) modelled them. If the weekday effects are removed from our model, it becomes equivalent to what S&E (2019) called the ‘Survivor Model’ with a calendar-time effect but without weekday effects. Figure S14 provides some support for this way of modelling the calendar-time effect, suggesting that it is indeed additive on the probit scale, at least for delays of two or more days. Whereas we apply the weekday effects to the hazard in equation (6), S&E’s (2020) model replaces equations (6) and (7) with

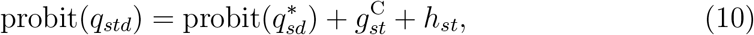

where *h_st_* is a cyclic cubic spline with a period of seven days (i.e. *h_st_* = *h_s,t_*_+7_). We believe that applying the weekday effects in equation (6) more accurately reflects these effects. Suppose, for example, that deaths are never reported on Sundays. This would be captured by our model through *η*_7_ = − ∞, but would not be captured by the spline *h_st_*, because *h_ts_* is a function of the weekday on which the individual died, rather than the weekday on which this death might be reported (see Appendix for further explanation). That weekday affects hazard of reporting is demonstrated by Figure 9, which shows the estimated distribution of delay by weekday of death. The probability of reporting within one day is approximately the same for deaths occurring on any of Monday to Friday, but is lower for deaths occurring on Saturday or Sunday. The probability of reporting within two days is the same for deaths occurring on Monday to Thursday, but lower for those occurring on Friday and Saturday (and to some extent Sunday). For reporting within three days, the probability is the same for deaths on Monday to Wednesday, but lower for Thursday to Saturday.

**Figure 9:**
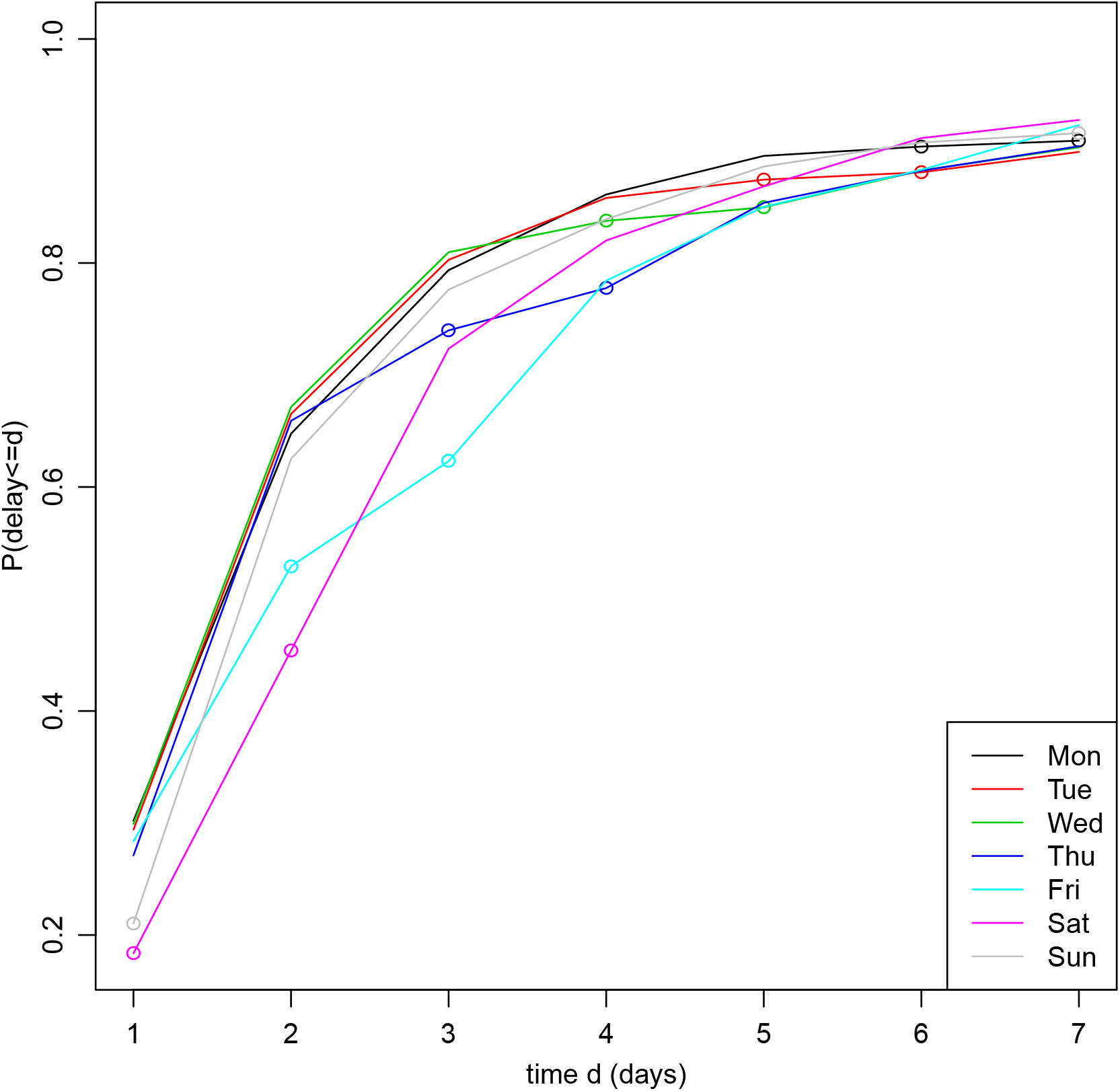
Estimated cumulative distribution function of delay given day of the week of death (only days 1–7 shown). The dots indicate Sundays and Mondays. (Estimate calculated using deaths that occurred at least 42 days before 29th June.)

An alternative way to model the calendar-time effect would be to apply it, together with the weekday effect, to the hazard in equation (6). This would yield what S&E (2019) call a ‘Hazard Model’. However, S&E (2019) argue that the Hazard Model formulation is less parsimonious than their Survival Model formulation (and hence our model), because it requires multiple splines for the calendar-time effect in each stratum: one for each possible delay *d*.

We used data from March 22nd onwards to estimate the delay distribution, with calendar-time effect splines and weekend effect splines to allow for change over time in this distribution. As data accumulates over a longer period of time, it may become desirable to use data only from the most recent period, e.g. the last *T* = 90 days. This would prevent the model-fitting computation time increasing over time and limit the danger that the calendar-time and weekend effect splines are insufficient to deal with changes over time in the delay distribution. To generate the results described in Section 3, which used *T* = 99, the computation time required to fit the model was 5.0 hours using one core of a MacBook Pro with a 2.6 GHz 6-core Intel i7 processor. This time was for the second adapted model of Section 2.3.4 and represents a reduction compared to that needed for the first adapted model (7.7 hours).

If computation time were a concern, two other measures that could be taken to reduce this, in addition to the measures already described, would be to decrease the maximum delay *D* and to increase the grouping of longer delays. We adjusted for delays of up to *D* = 42 days. S&E adjusted for delays of up to only *D* = 14 days. Overall, around 97% of deaths that are reported within 42 days are reported within 14 days, and so this restriction may be reasonable. We distinguished between delays of *d* and *d* + 1 days all the way up to *d* = 14 days, grouping longer delays into weeks. S&E, on the other hand, grouped together all delays of more than seven days. This reduces the computation time, but has the drawback that some information is discarded. Specifically, data on deaths that occurred between 8 and 13 days ago and that have been reported with delays of more than seven days are ignored. As well as potentially reducing statistical efficiency, this discarding of information allows the estimate of the number of deaths that occurred on any of these days to be lower than the number so far observed to have occurred on that day. It was for these reasons that we waited until 15 days before grouping delays and thereafter grouped them into weeks rather than into a single group.

The definition of a CoVID-19 related death is changing, with a number of alternatives being proposed based on the interval between the date of first positive test and the death event (https://assets.publishing.service.gov.uk/government/uploads/system/uploads/attachment_data/file/908781/Technical_Summary_PHE_Data_Series_COVID-19_Deaths_20200812.pdf). Our approach would still be appropriate to look at trends in these new data.

Finally, delayed reporting is a feature of administrative datasets, so the same problem would also affect the reporting of new CoVID-19 positive test from the current pillar schemes (https://assets.publishing.service.gov.uk/government/uploads/system/uploads/attachment_data/file/878121/coronavirus-covid-19-testing-strategy.pdf). Adjustment for reporting delays of these data is even more crucial to a prompt monitoring of the pandemic. Although it could be addressed through the approaches presented here, further modelling considerations as well as computational efficiency would be required.

## Data Availability

All data used in the analyses of this paper are daily totals aggregated to the seven NHS Regions and age-groups of broad width. These data are available upon the signing of a data-sharing agreement with Public Health England.

## Acknowledgements

This work was supported by the Medical Research Council (Unit programme numbers MC UU 00002/10 and MC UU 00002/11) in partnership with Public Health England. We acknowledge the support of the PHE Epidemiology Cell in consistently providing the data streams used.

## Appendix

### Likelihood of second adapted model in Section 2.3.4

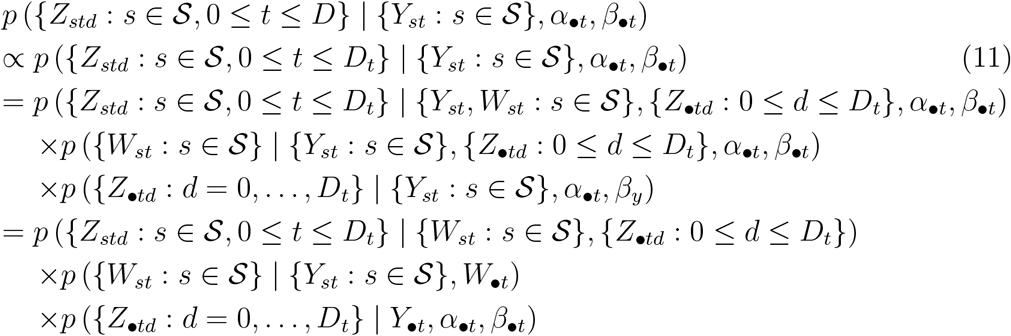

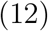

The proportionality sign in expression (11) reflects the fact that *Z_std_* is unobserved when *d* > *D_t_*, and so can be integrated out (as mentioned in Section 2.2). The first term in expression (12) is the likelihood of a generalised hypergeometric distribution [7]. It is a function only of observed variables, and hence can be ignored when calculating full-conditional distributions in the MCMC algorithm.

### Weekday effect example in Section 4

For simplicity, we ignore 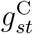 and 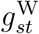 and omit the subscript *s* for stratum. Suppose deaths are never reported on Sundays but are reported on all other days of the week, and suppose day *t* is a Sunday. For someone who died on day *t*, the probability of being reported on day *t* equals zero, and hence *q_t_*_0_ equals zero, but their (expected) probability *q_t_*_1_ of being reported by the end of the following day would be non-zero, say *q_t_*_1_ = *c_t_*. So, from equation (10) we have that 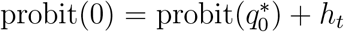 and 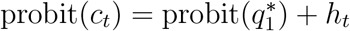. Since c_t_ > 0, it follows that 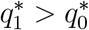.

On the other hand, for someone who died on day *t* − 1, which was Saturday, the (expected) probability q_t-1_,_0_ of being reported on day *t* − 1 would be non-zero, say *q_t_*_−1_,_0_ = c_t-1_, and their probability of being reported by the end of the following day would be no greater, i.e. *q_t_*_−1_,_1_ = *q_t_*_−1_,_0_ = *c_t_*_−1_. So, from equation (10) we have that 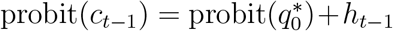 and 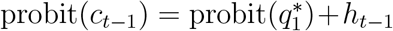. It follows from this that 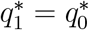, which contradicts our earlier deduction that 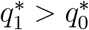.

